# Metagenomic Profiling of Airborne Microbial Communities from Aircraft Filters and Face Masks

**DOI:** 10.1101/2025.02.26.25322977

**Authors:** Oliyad Jeilu, Jack T. Sumner, Anahid A. Moghadam, Kelsey N. Thompson, Curtis Huttenhower, Charlie Catlett, Erica M. Hartmann

## Abstract

Airborne microbial communities, although often challenging to study due to low biomass, play crucial roles in public health and pathogen transmission. Through shotgun metagenomics, this study utilizes non-invasive air sampling of face masks and aircraft cabin filters to investigate microbial diversity in environments with frequent human interactions, including hospitals and airplanes. A comprehensive sampling and analysis workflow was developed, incorporating environmental and enrichment protocols to enhance microbial DNA recovery and diversity profiling. Despite limitations in biomass, optimized extraction methods allowed for the successful identification of 407 species, with dominant taxa including *Cutibacterium acnes*, *Staphylococcus epidermidis*, *Sphingomonas hankookensis*, and *Methylobacterium radiotolerans*. Enrichment processing resulted in greater metagenome-assembled genome (MAG) recovery and higher antimicrobial resistance gene (ARG) identification. The findings highlight the presence of ARGs in high-occupancy public spaces, suggesting the importance of monitoring and the potential for mitigating airborne transmission risks in such environments. This study demonstrates the utility of combining environmental and enrichment sampling to capture comprehensive microbial and ARG profiles in confined spaces, providing a framework for enhanced pathogen monitoring in public health contexts.

## Background

Although often seen as inhospitable for microbial life due to its low nutrient availability, air is a critical medium for the transport and dispersal of microorganisms [1,2]. These airborne microbes originate from various natural sources, such as soil, oceans, vegetation, and anthropogenic activities [3]. While these microorganisms play roles in ecological processes, airborne transport may significantly impact human health by transmitting infectious pathogens [4,5].

The study of airborne microbes has historically lagged research on other environments, such as soil and water, mainly due to the challenges in sampling and analyzing these microorganisms [7,8]. However, the COVID-19 pandemic, fueled by airborne transmission, led to a surge in research on airborne microbes [6,9]. While culture-based studies have been foundational, less than 1% of airborne prokaryotes are cultivable using traditional microbiological procedures, offering an incomplete picture of microbial community structures and dynamics [10–12].

In recent years, culture-independent techniques, particularly metagenomics, have emerged as powerful tools for studying the structure and function of airborne microbial communities and pathogens [11,13]. Metagenomics provides a broader assessment of microbial diversity and metabolic potential than culture-based techniques. However, the application of metagenomics to airborne microbial research presents challenges. Low biomass, sample collection logistics, DNA extraction difficulties, and contamination issues have hindered comprehensive analysis of airborne microbial communities [8,14,15]. These technical challenges limit the detection of low-abundance organisms and can complicate efforts to generate reliable data.

In high-risk environments such as aircraft cabins and hospitals, face masks and ventilation systems equipped with High-Efficiency Particulate Air (HEPA) filters play crucial roles in reducing the transmission of airborne pathogens [16,17]. HEPA filters capture 99.97% of airborne particles, including bacteria and viruses, but do not destroy trapped microbes, allowing viable ones to multiply and pose a recontamination risk [18].

Similarly, face masks block infectious droplets and aerosols, but improper handling, like frequent hand contact, can lead to microbial buildup, reducing their protective effectiveness [19]. While potentially problematic for proper use and maintenance, the ability of HEPA filters and face masks to trap and retain microorganisms may incidentally provide a convenient mechanism for air sampling.

During a pandemic, it would be valuable to determine whether an airborne pathogen is present in various situations, e.g., on flights or within a healthcare environment. Air handling systems provide an opportunity to sample at the population level for the enclosed area, hence our initial interest in aircraft cabin air filters. However, given that these are typically expensive to change after 8-10,000 flight hours, analyzing disposable face masks seems a more logistically realistic strategy.

This study explores the diversity and function of airborne microbial communities through shotgun metagenomics, focusing on samples collected from face masks worn by air travelers and physicians in hospitals and an aircraft HEPA filter. Given the low biomass of airborne microbial samples, we optimized DNA extraction protocols and incorporated enrichment protocols using Brain Heart Infusion (BHI) media to stimulate the growth of airborne microorganisms. This research seeks to bridge critical gaps in understanding microbial populations in confined environments while evaluating approaches that may inform public health risk assessment.

## Methodology

### Sample Collection and Processing

One aircraft cabin filter was removed from a commercial airplane after 8,039 hours of usage. The filter was immediately placed in a sterile, DNA-free plastic bag and transported to the laboratory. In an aseptic environment, the filter was manually disassembled, and the filtration material was carefully removed using sterilized, DNA-free scissors. The filter material was then placed into sterile, DNA-free plastic bags and stored in a refrigerator until DNA extraction to minimize contamination and preserve sample integrity. Samples were taken randomly from different areas of the filter. Each sample measured approximately 4 cm by 4 cm.

Face masks worn by air travelers and healthcare professionals in a hospital unit or ward were collected after a single use. Each sample was placed in a sterile bag and transported to the laboratory. Unworn face masks exposed to environmental conditions of an aircraft during flight were also collected.

The outer layer of each collected face mask and aircraft cabin filter sample was sectioned into 4 cm by 4 cm pieces using sterile scissors. These were divided into two parts; one 2 cm by 4 cm portion was used for direct DNA extraction, and the other half was used for enrichment before DNA extraction.

### Enrichment Culture

Samples from facemasks and the aircraft cabin air filter were incubated in 50 ml Brain Heart Infusion media at 37°C for up to 5 days with agitation at 120 rpm. Aliquots of 1 ml were taken every 24 hours, and microbial cells were collected by centrifugation. The extracted DNA from these aliquots, originating from the same sample, was pooled together for downstream analysis. These samples are hereafter referred to as “Enrichment,” and unenriched samples are called “Environmental.”

### DNA Extraction

DNA extraction was performed using the optimized method described by [15] and utilizing the DNeasy PowerSoil Kit (Qiagen, Hilden, Germany). The sectioned samples were placed into PowerBead tubes supplied with the kit. Lysis buffer was added at twice the volume recommended by the manufacturer. The tubes were incubated at 37°C for 30 minutes to enhance lysis efficiency. Following incubation, the samples were homogenized at a lower speed of 200 rpm for 1 hour and 30 minutes to mix the contents gently. After homogenization, the samples were centrifuged at a medium speed of 2,000 x g for 10 minutes to pellet debris while preserving the integrity of the DNA. The supernatant was then carefully transferred to a new tube, ensuring no debris was carried over into the final sample. The extraction process was then continued according to the standard protocol provided with the DNeasy PowerSoil Kit, including steps for chemical precipitation, washing, and DNA elution. The concentration and quality of the extracted DNA were initially assessed using a Qubit 4 fluorometer with the dsDNA HS assay kit (Thermo Fisher Scientific, Waltham, MA, USA).

### PCR Amplification of 16S rRNA Gene and Quantitative PCR (qPCR) for Microbial DNA Quantification

Due to the low biomass nature of the samples, the DNA concentrations were often below the detection limit of the Qubit system. Therefore, PCR amplification of the 16S rRNA gene was performed to confirm the presence of microbial DNA using the standard primers 515F (5’-GTGCCAGCMGCCGCGGTAA-3’) and 806R (5’-GGACTACHVGGGTWTCTAAT-3’) [20]. The reaction was carried out in a 25 µL volume using Promega Green GoTaq™ master mix (Promega Corporation, Madison, WI, USA) containing 1X Green GoTaq buffer, 0.2 µM of each primer, and 5 µL of template DNA. The thermal cycling conditions were as follows: an initial denaturation at 95°C for 3 minutes, followed by 35 cycles of denaturation at 95°C for 30 seconds, annealing at 55°C for 30 seconds, and extension at 72°C for 30 seconds, with a final extension at 72°C for 5 minutes. PCR products were analyzed by agarose gel electrophoresis to determine successful amplification and the expected product size.

Quantitative real-time PCR (qPCR) was performed to quantify microbial 16S rRNA gene copies using PowerUp™ SYBR™ Green Master Mix (Thermo Fisher Scientific, Waltham, MA, USA) on a QuantStudio™ Real-Time PCR System (Thermo Fisher Scientific). Each reaction was performed in a total volume of 20 µL, containing 10 µL of 2X PowerUp™ SYBR™ Green Master Mix, 1 µL of each 10 µM primer (515F and 806R), 2 µL of template DNA, and 6 µL of nuclease-free water. The thermal cycling protocol included an initial uracil-DNA glycosylase (UDG) activation step at 50°C for 2 minutes to remove carryover contamination, followed by Dual-Lock™ DNA polymerase activation at 95°C for 2 minutes.

Amplification was performed over 40 cycles of denaturation at 95°C for 15 seconds and annealing/extension at 60°C for 30 seconds. A melt curve analysis was conducted with denaturation at 95°C for 1 second, annealing at 60°C for 20 seconds, and a gradual increase in temperature to 95°C at a rate of 0.1°C per second with continuous fluorescence measurement.

Quantification of 16S rRNA gene copies was achieved by generating a standard curve using serial tenfold dilutions of a 4,125 bp *Escherichia coli* plasmid containing a 16S rRNA gene insert [21]. The standard curve had an R² value of 0.96, and the qPCR efficiency was calculated to be 111%. All qPCR reactions were performed in triplicate, and a no-template control (NTC) was included in each run to monitor for contamination or non-specific amplification. Normalization was performed using a microbial spike-in control to account for variability in DNA recovery and technical inconsistencies during sample processing. A defined quantity of spike-in DNA (20 µL at a theoretical concentration of 10⁹ copies/µL, equivalent to 2 × 10¹⁰ copies) was added to each sample before DNA extraction. The observed spike DNA copy number was quantified in each sample via qPCR, and the recovery efficiency of the spike was calculated as the ratio of observed spike copies to the theoretical spike copies (5.27 × 10⁻⁷). This recovery efficiency was then used to adjust each sample’s 16S rRNA gene copy numbers, ensuring accurate quantification that accounts for potential losses during DNA extraction or other processing steps.

### Spike-In Controls

Spike control samples were prepared by inoculating a 2 cm x 2 cm sectioned sterile face mask with the ZymoBIOMICS Microbial Community Standard (Zymo Research, Irvine, CA, USA; Catalogue No. D6300). Each section was inoculated with 100 µL of the ZymoBIOMICS Microbial Community Standard. The inoculated face mask sections were incubated at 37°C for 30 minutes to ensure microbial adhesion. Following the incubation, the DNA extraction was performed as mentioned above. The standard contains a defined mixture of microbial species with the following theoretical composition based on genomic DNA: *Listeria monocytogenes* - 12%, *Pseudomonas aeruginosa* - 12%, *Bacillus subtilis* - 12%, *Escherichia coli* - 12%, *Salmonella enterica* - 12%, *Lactobacillus fermentum* - 12%, *Enterococcus faecalis* - 12%, *Staphylococcus aureus* - 12%, *Saccharomyces cerevisiae* - 2%, and *Cryptococcus neoformans* - 2%.

### Shotgun Metagenomic Sequencing

Shotgun metagenomic sequencing was performed using the Illumina NextSeq 2000 platform, employing a 300-cycle flow cell kit to produce 2 × 150 bp paired-end reads. The Illumina DNA Prep (tagmentation) kit prepared libraries with unique dual indexes. PCR cycles were increased to accommodate the low DNA input, following the manufacturer’s protocol for low-input library preparation. A 1–2% PhiX control was spiked into the sequencing run to support optimal base calling. Sequencing was performed to achieve a targeted depth of 1000 Mbp, corresponding to approximately 6.7 million paired-end reads per sample. The sequencing process ensured comprehensive coverage suitable for detecting low-abundance taxa in metagenomic samples.

### Bioinformatics Analysis

#### Quality Control and Contaminant Removal

Raw sequencing reads were first assessed using FastQC v 0.12.0 [22], and then, sequencing adapters and low-quality bases were trimmed using Trimmomatic v 0.39 ([23] with specific parameters (SLIDINGWINDOW:4:20 MINLEN:50) to retain only high-quality reads. Following KneadData (https://huttenhower.sph.harvard.edu/kneaddata/) default parameters, the human genome DNA and PhiX control DNA sequences were filtered out. Additionally, overrepresented sequences were identified and removed. Finally, the clean reads underwent a final round of quality assessment using FastQC v 0.12.0, and the results were compiled with MultiQC v 1.23 [24] to generate a comprehensive summary report.

#### Read-Based Microbial Community Profiling and Functional Analysis

Taxonomic profiling of purified metagenomic reads was performed using MetaPhlAn4 [25], utilizing the Jun23_CHOCOPhlAnSGB_202307 database of marker genes. Subsequent analyses of the MetaPhlAn output included evaluations of microbial community structure and the calculation of diversity indices.

Strain-level resolution was achieved using StrainPhlAn 4.1 [26], focusing on three abundant species across the samples: *Cutibacterium acnes* (t SGB16955), *Staphylococcus epidermidis* (t SGB7865), and *Staphylococcus hominis* (t SGB7858).

Functional profiling was conducted using HUMAnN 3 [27], utilizing the ChocoPhlAn database (v201901_v31) for taxonomic gene family annotations and the UniRef50 database (uniref50_201901b) for protein functional assignments. Following functional profiling, gene families were regrouped into MetaCyc reactions using predefined mapping files provided by HUMAnN. After regrouping, the data were normalized to relative abundance. The non-stratified table, which compiles pathway data from all species, was employed for further downstream analyses and visualizations.

#### Diversity Analysis

Diversity analyses were performed on microbial community profiles generated by MetaPhlAn. Alpha diversity, measured by the Shannon index, was calculated for each sample using species-level abundance data. Statistical significance between sample types was tested using the Kruskal-Wallis test, followed by pairwise Wilcoxon rank-sum tests with Benjamini-Hochberg correction. Bray-Curtis dissimilarity matrices were computed from MetaPhlAn’s species abundance profiles for beta diversity. Principal Coordinate Analysis (PCoA) was performed to visualize differences in community composition across sample types. PERMANOVA analysis, as implemented in the adonis2 function of the vegan package v 2.6-4 [28], was used to statistically assess variation in community structure between sample types, with p-values included in the PCoA plots.

#### Metagenomic Assembly, Genome Construction, and Coverage Analysis

The clean metagenomic reads were assembled into contigs using MEGAHIT v1.2.9 [29], using the default parameters. After assembly, the quality of the resulting contigs was evaluated using QUAST v5.2.0 [30]. The contigs were processed into metagenome-assembled genomes (MAGs) via MetaBAT 2 [31]. The binning procedure was initiated by aligning the raw paired-end reads to the assembled contigs using bowtie2 v2.5.4 [32]. These aligned reads were sorted and indexed using Samtools v1.20 [33], generating the BAM files necessary for MetaBAT 2. MetaBAT 2 was then run with its default parameters, utilizing these BAM files alongside the contig assemblies to delineate MAGs. After binning, the quality of the MAGs was evaluated using CheckM v1.1.6 [34]. Each bin generated from MetaBAT 2 was analyzed with CheckM to determine the completeness and contamination levels of the assembled genomes. The MAGs with a completeness of 50% or greater and contamination of 10% or less were retained for downstream analysis.

Log-transformed coverage values were visualized to represent the distribution and density of coverage to analyze differences in coverage between enrichment and environmental processing conditions. Kruskal-Wallis tests were used to assess statistical differences in microbial coverage between sample types, and outliers, identified as points beyond 1.5 times the interquartile range (IQR), were highlighted as red circles in the plots. Coverage comparisons between Enrichment and Environmental conditions were performed using Wilcoxon signed-rank tests, and significant differences were annotated on the plots.

#### Taxonomic and Functional Classification of the MAGs

MAGs underwent taxonomic classification using GTDB-Tk v2.4.0 [35] performed using the most recent GTDB release (Release 220). Prokka v1.14.5 [36] was employed to annotate high-quality MAGs. Antibiotic resistance gene identification within the MAGs was performed using the CARD (Comprehensive Antibiotic Resistance Database) [37] via the Abricate tool v1.0.0 [38].

#### Comparative Analysis of Microbial Diversity

A comparative analysis between species detected by MetaPhlAn and GTDB-Tk was performed to identify the overlap in species identification and highlight the species exclusively identified by either tool. Species presence and relative abundances were compared across the Environmental and Enrichment processing using the Wilcoxon signed-rank test for paired samples. The test evaluated significant differences in species abundance between Environmental and Enrichment samples.

## Results

### Microbial Composition of Face Masks and Aircraft Cabin Filters

We collected one aircraft cabin air filter, 29 face masks worn by travelers, six unworn face masks exposed to aircraft cabin conditions, and 18 worn by healthcare professionals. A total of 55 samples were successfully sequenced, comprising 19 hospital samples, 12 aircraft filter samples (from a single Aircraft filter cabinet), 16 traveler samples, and eight control samples (six Unworn and two spike controls). These samples were processed using two different methods: 22 Enrichment samples and 31 Environmental samples, along with the two spike control samples. Environmental samples showed lower DNA recovery (∼0.48 ng/µL) and higher Ct values (∼17.36), with limited variability across sample types. In contrast, Enrichment samples consistently yielded the highest DNA concentrations (∼20.64 ng/µL), lower Ct values (∼15.62), and higher copy numbers (∼10^16^). Among Enrichment samples, Hospital settings exhibited the highest DNA concentrations (∼45.44 ng/µL) and copy numbers (∼10^16^), while Travel samples showed the lowest Ct values (∼12.82), indicating efficient amplification despite lower DNA concentrations. Aircraft Filter samples under Environmental processing demonstrated the lowest DNA recovery, highlighting challenges in low-biomass sample types (Figure 1).

**Figure 1:**
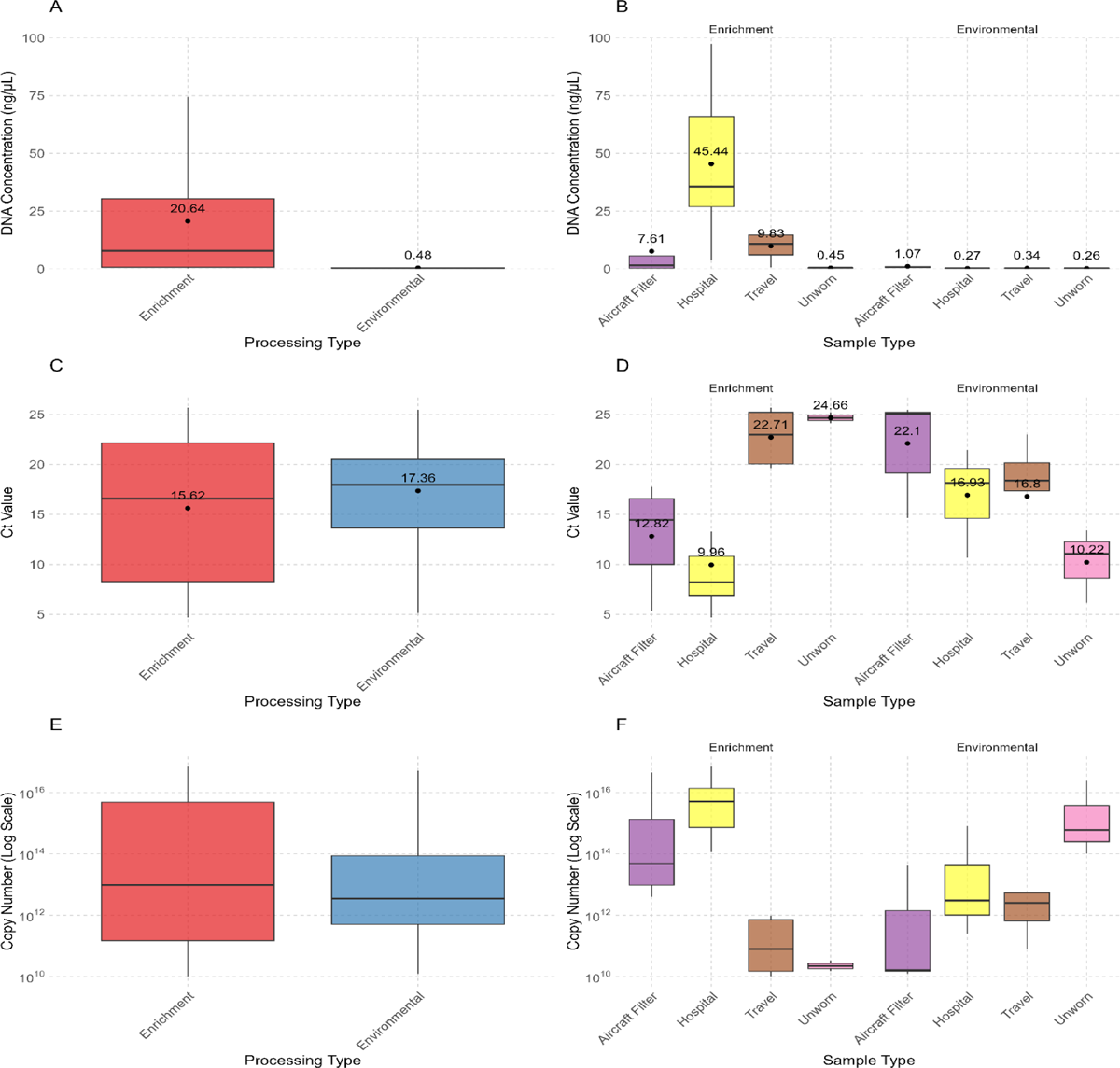
Comparative Analysis of DNA Concentration, Ct Values, and Copy Numbers. DNA concentration (ng/µL) (A) across processing types and (B) across sample types within each processing type. Ct values (C) across processing types and (D) across sample types within each processing type. Copy numbers (copy number/µl) on a logarithmic scale (E) across processing and (F) across sample types within each processing type. Mean values are marked with black dots and annotated above the boxes.

Sequencing of these samples generated a total of 1,677,589,656 raw reads. After quality control, the cleaned dataset consisted of 1,279,623,449 reads, representing a 23.7% reduction in the total number of reads (Supplementary Table 1).

The microbial community composition analysis at the domain level revealed that most sequences were classified as Bacteria, accounting for 99.6% of the total relative abundance. A small fraction of the community was classified as Eukaryota, contributing 0.446% to the relative abundance. Further taxonomic breakdown identified 12 distinct phyla and 407 species across the samples (Table 1).

**Table 1:** Relative Abundance of Phyla within Bacteria and Eukaryota Kingdoms.

| Kingdom | Phylum | Relative abundance (%) |  |
| --- | --- | --- | --- |
|  |  | Domain | Total |
| **Bacteria** | Acidobacteria | 0.002 | 0.002 |
| **Bacteria** | Actinobacteria | 32.6 | 32.46 |
| **Bacteria** | Bacteroidota | 1.5 | 1.5 |
| **Bacteria** | Candidatus_Absconditabacteria | 0.01 | 0.009 |
| **Bacteria** | Candidatus_Saccharibacteria | 0.25 | 0.25 |
| **Bacteria** | Deinococcus_Thermus | 0.07 | 0.06 |
| **Bacteria** | Firmicutes | 40.0 | 39.9 |
| **Bacteria** | Fusobacteria | 0.05 | 0.05 |
| **Bacteria** | Proteobacteria | 12.9 | 12.8 |
| **Bacteria** | Spirochaetes | 0.001 | 0.001 |
| **Bacteria** | Unclassified | 12.5 | 12.5 |
| **Eukaryota** | Ascomycota | 2.3 | 0.01 |
| **Eukaryota** | Basidiomycota | 85.2 | 0.38 |
| **Eukaryota** | Unclassified | 12.5 | 0.056 |

The phylum level revealed notable differences in relative abundance between Environmental and Enrichment processing methods across the sample types. Actinobacteria exhibited a substantial presence across all sample types and processing methods, with an exceptionally high relative abundance in environmental samples, 41.5% in Travel samples, and 32.5% in Hospital samples. Firmicutes showed a significant presence in Enrichment samples, especially in Travel (29.5%) and Hospital (29.0%) samples, whereas its relative abundance in Environmental samples was notably lower (Figure 2B). Other phyla, such as Bacteroidota, Candidatus Saccharibacteria, and Fusobacteria, exhibited lower relative abundances, generally below 1%, across all sample types and processing methods (Figure 2A-B).

**Figure 2:**
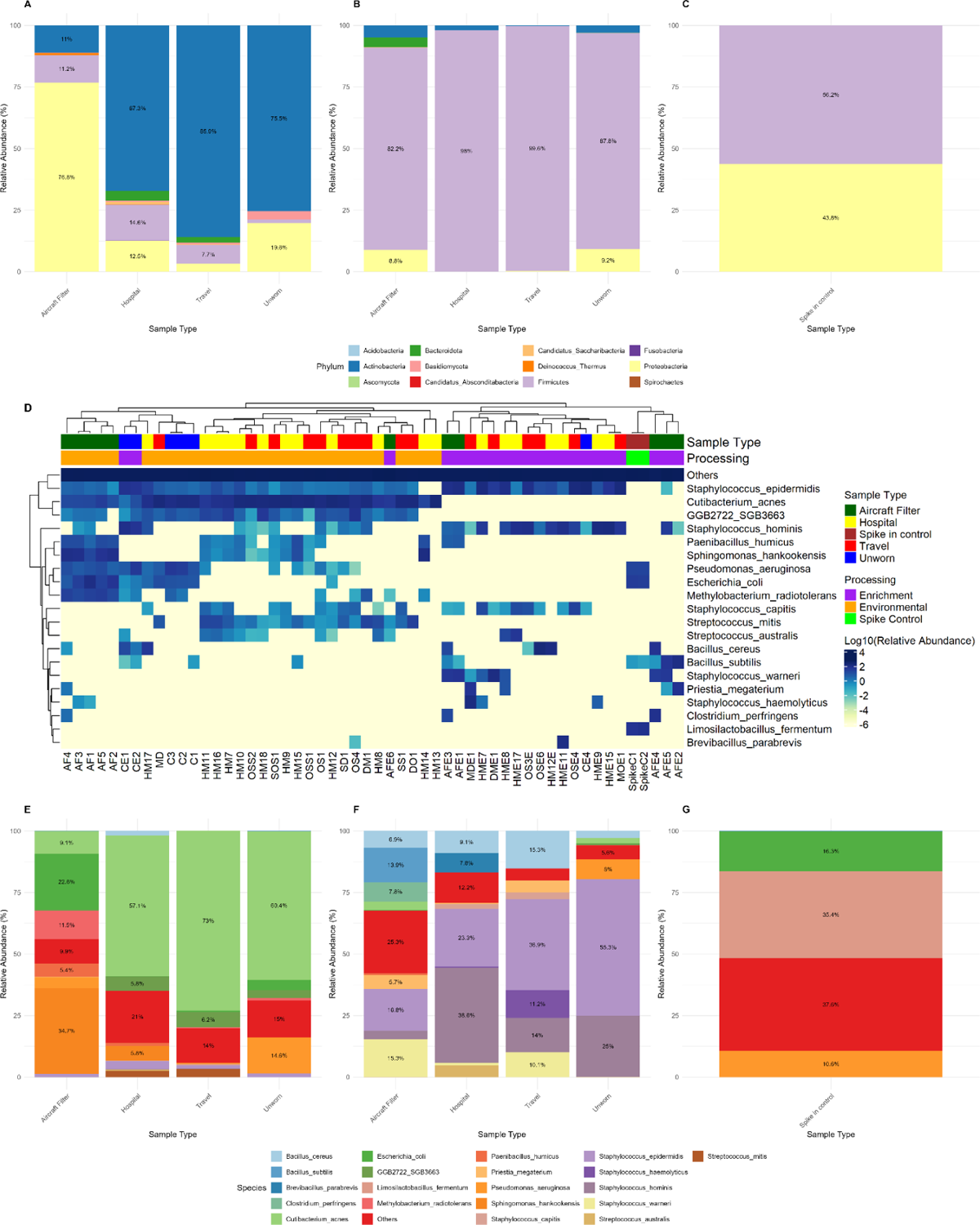
Microbial community composition across different sample types and processing methods. Panels (A–C) represent the relative abundances of microbial phyla in environmental, enrichment, and spike control samples, respectively. Panel (D) shows a heatmap of the log10-transformed relative abundances of the top 20 most abundant species across all samples, with metadata annotations for sample type and processing method displayed above the heatmap. Panels (E–G) depict species-level relative abundances in environmental, enrichment, and spike control samples. For the barplots, the labels highlight taxa with >5% abundance.

*Cutibacterium acnes*, *Staphylococcus epidermidis*, and *Staphylococcus hominis* emerged as the dominant species across many samples. Other species, including *E. coli* and *Sphingomonas hankookensis*, also contributed to the microbial diversity, albeit with varying abundance levels depending on the sample type and processing method (Figure 2D).

In Environmental samples, *C. acnes* were highly dominant, especially in Travel and Hospital samples, with relative abundances reaching 73.0% and 57.1%, respectively. In the Aircraft Filter samples, *S. hankookensis* emerged as the most abundant species, constituting 34.7 % of the microbial community, followed by *E. coli* at 22.6 % (Figure 2E). Under Enrichment conditions, there was a shift in the microbial community structure across different sample types, with *S. epidermidis* emerging as a particularly dominant species. Furthermore, in Hospital samples, *S. hominis* also became prominent under Enrichment conditions, with a relative abundance of 38.6 % (Figure 2F).

The phylogenetic tree of *C. acnes* (Figure 3A), *S. epidermidis* (Figure 3B), and *S. hominis* (Figure 3C) reveals significant strain-level diversity. These strains from various sample types, including Hospital, Travel, Aircraft Filter, and Unworn, are well distributed across the tree.

**Figure 3:**
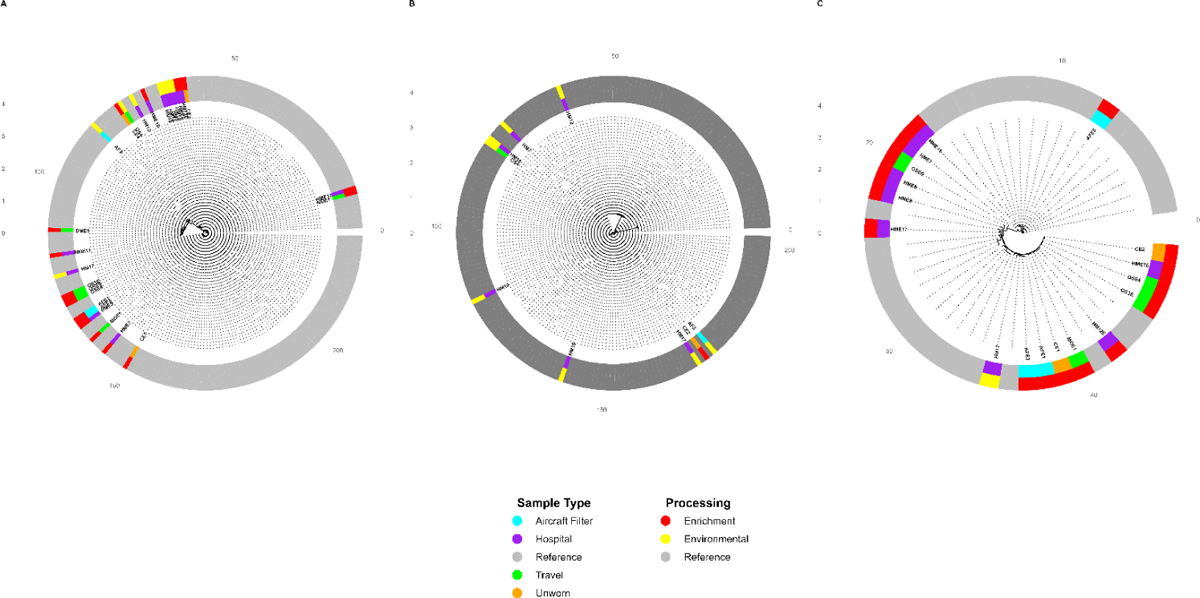
Circular phylogenetic trees of three bacterial species derived from StrainPhlAn analysis. (A) *Staphylococcus epidermidis* (t SGB7865), (B) *Cutibacterium acnes* (t SGB16955), (C) *Staphylococcus hominis* (t SGB7858). Each branch represents a unique strain, with color-coded rings indicating the sample type and processing.

### Microbial Diversity and Spatial Distribution

The Shannon diversity index varied across different sample types under environmental and enrichment conditions. Environmental samples demonstrated distinct diversity patterns (Figure 4A). Hospital samples exhibited the highest Shannon diversity, followed by Aircraft Filter samples, which showed a broader range of values. In contrast, Travel-related samples had the lowest median diversity (Figure 4B). Under enrichment conditions, Travel-related samples consistently exhibited the lowest diversity, mirroring trends observed under environmental conditions. Significant diversity differences were observed across Aircraft filter and Travel samples (p < 0.05, Figure 4C).

**Figure 4:**
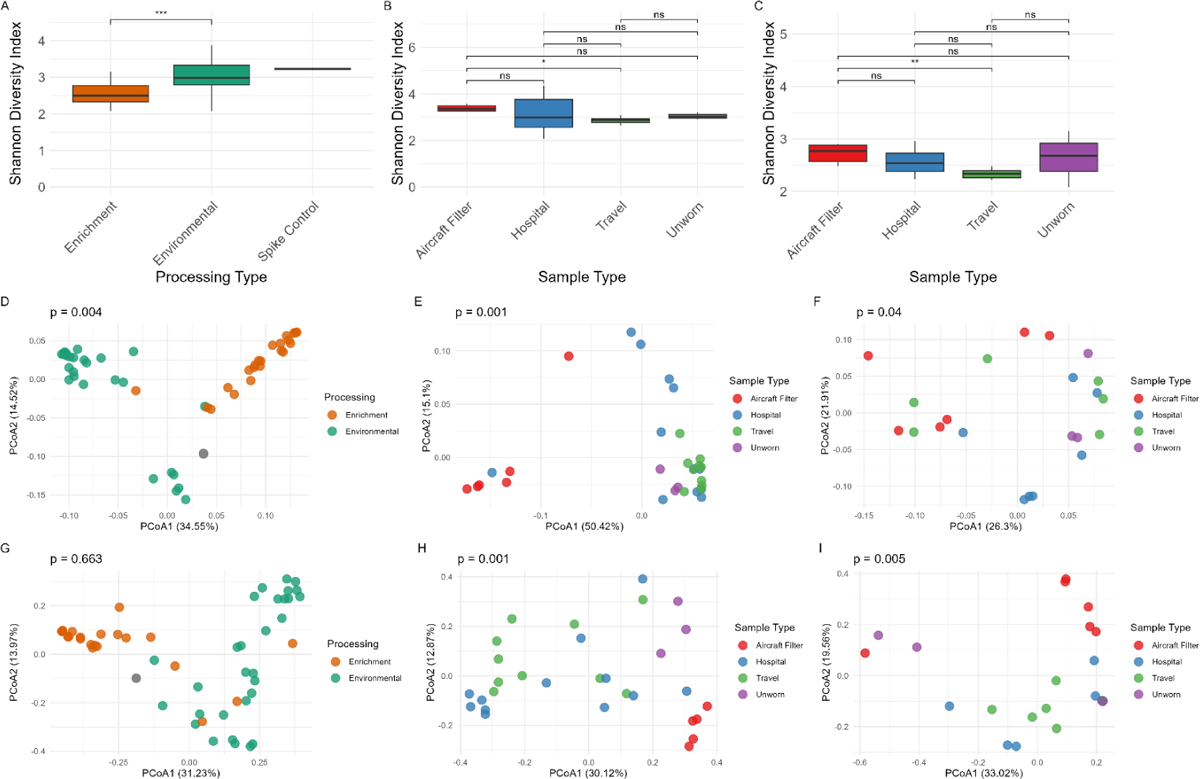
Alpha and beta diversity of microbial communities across different sample types and processing methods. The Shannon Diversity Index is shown by sample type for (A) Environmental vs. Enrichment, (B) Environmental samples, and (C) Enrichment samples. Beta diversity analyses based on unweighted UniFrac distances are displayed for (D) Environmental vs. Enrichment, (E) Environmental samples, and (F) Enrichment samples. Beta diversity analyses based on Bray-Curtis dissimilarity are shown for (G) Environmental vs. Enrichment, (H) Environmental samples and (I) Enrichment samples.

**Figure 4:**
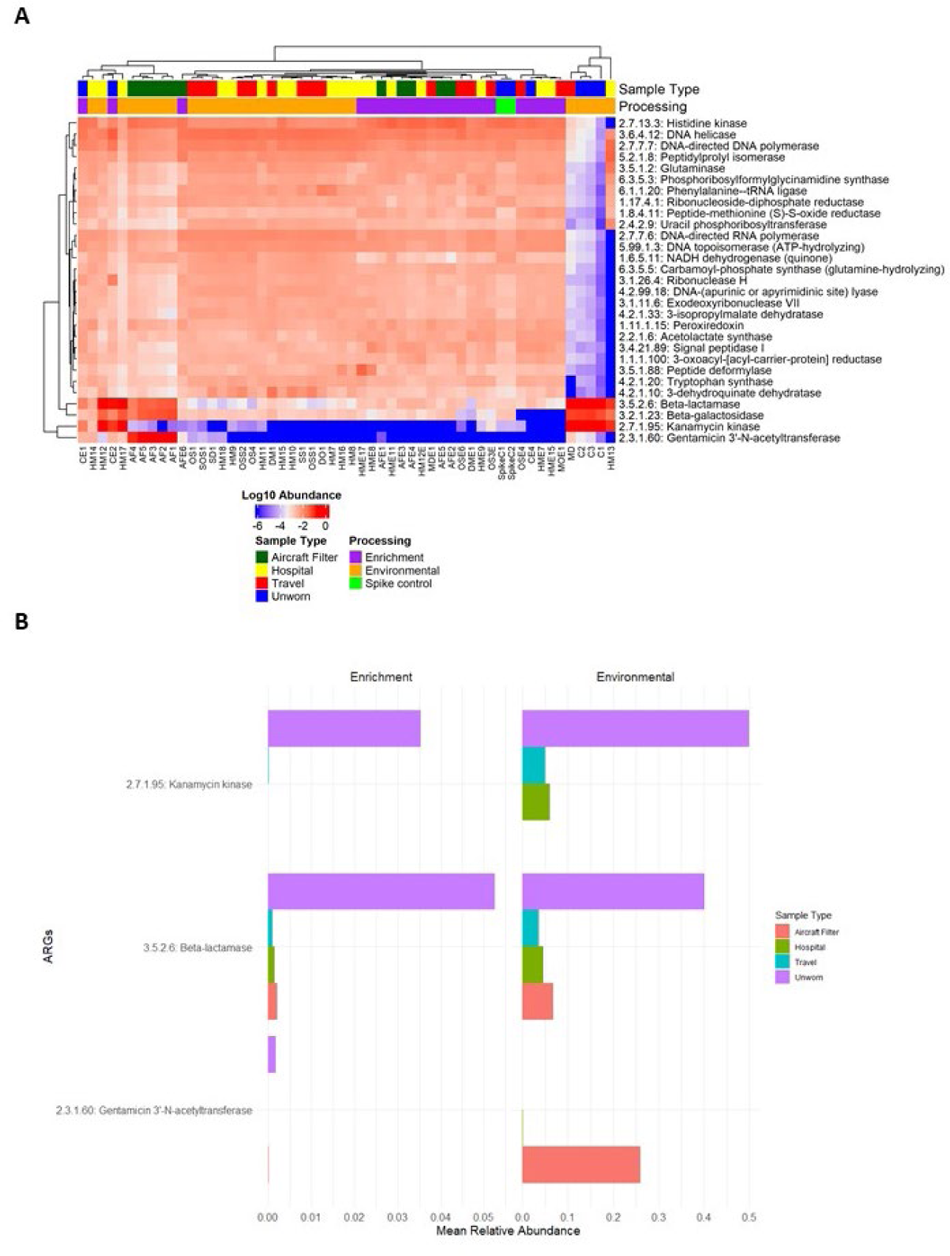
Top 30 Gene Families from HUMAnN3 MetaCyc Pathway and Antibiotic Resistance Gene Abundance Across Sample Types and Processing Categories. (A) Heatmap displaying the top 30 gene families identified, shown as Log10 relative activity. (B) Stacked bar plot showing the mean relative abundance of the antibiotic resistance gene families.

Beta diversity analysis based on unweighted UniFrac distances revealed no significant separation between environmental and enrichment samples (PERMANOVA p = 0.652, Figure 4D). Within environmental samples, distinct clustering was observed among sample types (PERMANOVA p = 0.013, Figure 4E). Aircraft Filter and Hospital samples formed tight clusters, while Travel-related samples were the most dispersed. Unworn samples partially overlapped with Hospital samples. For enrichment samples, clustering patterns were also evident (PERMANOVA p = 0.046, Figure 4F).

Analysis using Bray-Curtis distances revealed significant separation between environmental and enrichment samples (PERMANOVA p = 0.001, Figure 4G). Within environmental samples, sample types clustered distinctly (PERMANOVA p = 0.022, Figure 4H), with tighter clustering observed for Hospital and Aircraft Filter samples compared to Travel-related and Unworn samples. For enrichment samples, distinct clustering patterns were also observed (PERMANOVA p = 0.012, Figure 4I). Aircraft Filter and Unworn samples exhibited tighter clusters, while Travel-related and Hospital samples were more dispersed.

### Functional Profiling of Gene Families Using HUMAnN3 MetaCyc Pathway Data

Gene families related to antimicrobial resistance, such as beta-lactamase (3.5.2.6), kanamycin kinase (2.7.1.95), and gentamicin 3’-N-acetyltransferase (2.3.1.60), were particularly enriched in Hospital samples compared to other sample types. In contrast, gene families involved in core metabolic functions, such as DNA-directed RNA polymerase (2.7.7.6) and glutaminase (3.5.1.2), exhibited a more even distribution across sample types (5A).

Notably, the abundance of ARGs was higher in Environmental samples than in Enrichment samples. Specifically, beta-lactamase and gentamicin 3-N-acetyltransferase were significantly more abundant in the Aircraft Filter and Unworn samples under Environmental samples. Conversely, kanamycin kinase showed a relatively higher abundance in the Unworn samples under Enrichment (Figure 5B).

**Figure 5:**
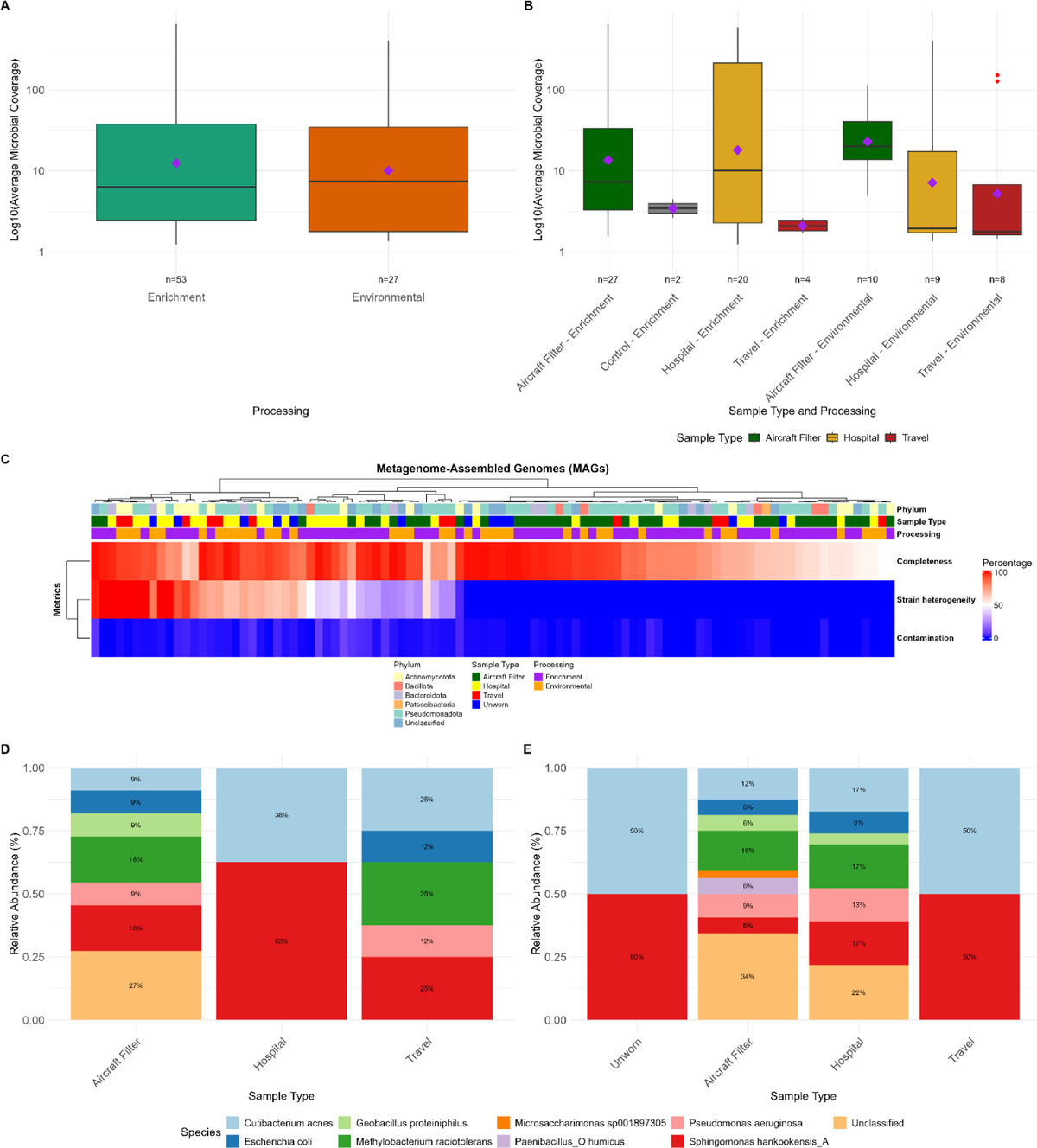
Box plots of average microbial coverage in different processing types and sample categories. (A) Comparison of average microbial coverage between Enrichment and Environmental processing types. (B) The breakdown of average microbial coverage by sample type and processing method. Mean values are marked by purple diamonds, with sample sizes indicated below each category. Outliers are represented as red points. (C) Heatmap of completeness, contamination, and strain heterogeneity metrics for MAGs and classified by GTDB-Tk at the phylum level. MAGs species-level relative abundance across different sample types under Environmental (D) and Enrichment (E) processing. Taxa with relative abundance greater than 5% are labeled with their corresponding percentages.

### Evaluation of Metagenome-Assembled Genomes

A total of 176 MAGs were recovered from various sample types and processing methods. The highest number of MAGs was obtained from Aircraft Filter samples processed through enrichment (65 MAGs), followed by Hospital samples processed through enrichment (43 MAGs). In terms of MAGs (having ≥50% completeness and ≤10% contamination), the Aircraft Filter samples processed under environmental conditions had a proportion of 44% (11 out of 25 MAGs), while those processed under enrichment conditions had a slightly higher proportion of 49.2% (32 out of 65 MAGs). No MAGs were recovered from the unworn Environmental samples. Hospital samples processed through environmental methods exhibited an average completeness of 84.9% and contamination of 0.8%, whereas those processed through enrichment had slightly higher average contamination at 1.9% but similar completeness (86.6%) (Table 2).

**Table 2:** Summary of metagenome-assembled genome metrics across different sample types and processing methods.

| Sample Type | Processing | Total MAGS | High-quality MAGS | Proportion | Average completeness | Average contamination |
| --- | --- | --- | --- | --- | --- | --- |
| Aircraft Filter | Enrichment | 65 | 32 | 49.2 | 79.7 | 1.2 |
| Aircraft Filter | Environmental | 25 | 11 | 44.0 | 86.6 | 0.5 |
| Unworn | Enrichment | 4 | 2 | 50.0 | 71.4 | 1.5 |
| Hospital | Enrichment | 43 | 22 | 51.2 | 86.6 | 1.9 |
| Hospital | Environmental | 14 | 8 | 57.1 | 84.9 | 0.8 |
| Travel | Enrichment | 8 | 4 | 50.0 | 81.6 | 1.4 |
| Travel | Environmental | 17 | 8 | 47.1 | 80.0 | 1.2 |

The analysis comparing the average microbial coverage between Enrichment and Environmental processing conditions revealed no statistically significant differences. The Enrichment samples, however, displayed a wider distribution of coverage values, with some samples exhibiting higher coverage, whereas the Environmental group showed a more compact distribution (Figure 6A). In the meantime, The Kruskal-Wallis test indicated significant differences in coverage among the sample types (*p* < 0.05), with Hospital and Aircraft Filter samples showing the highest coverage under both Enrichment and Environmental conditions (Figure 6B).

**Figure 6:**
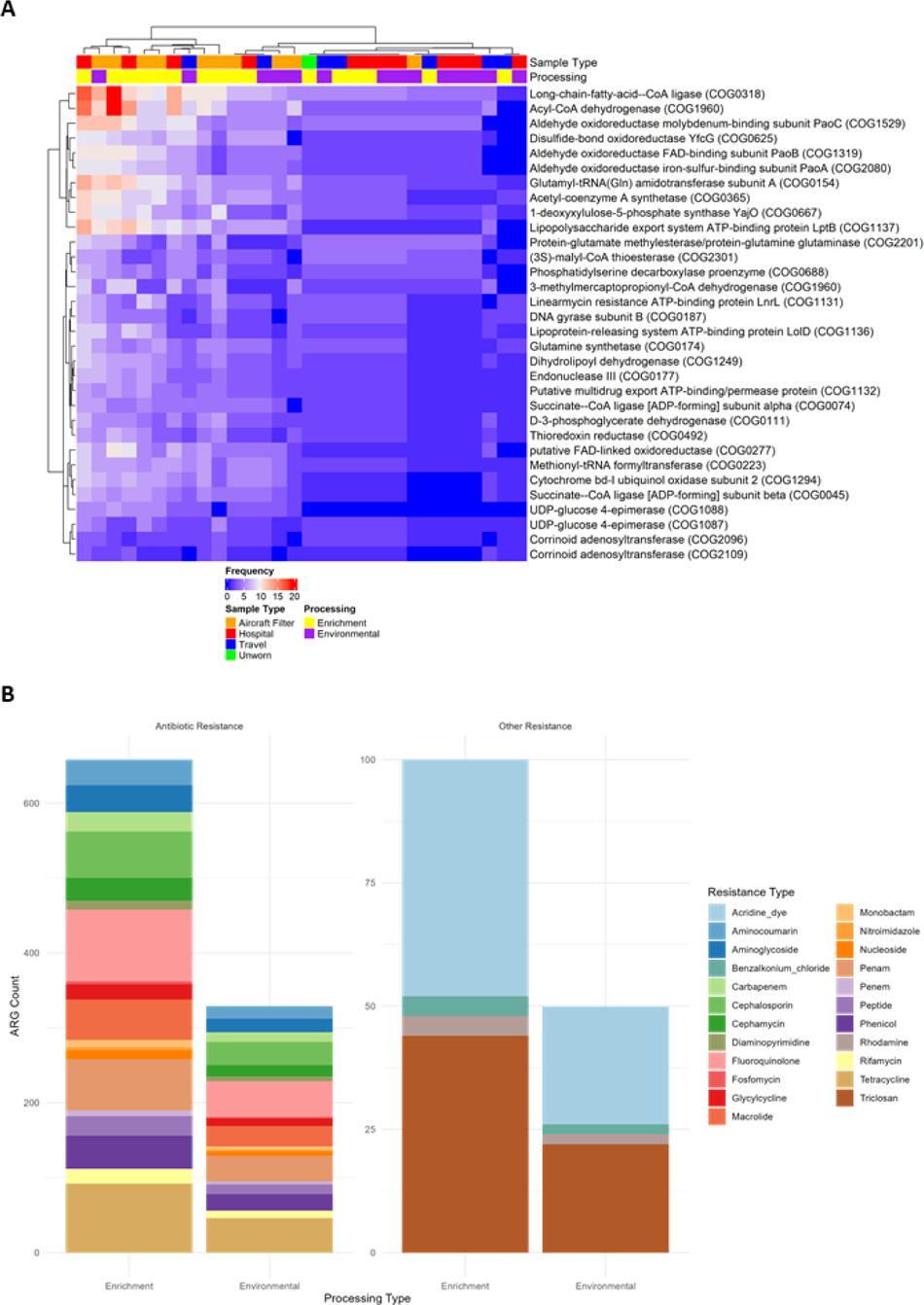
(A) Heatmaps of Top 30 Gene Products in both Environmental and Enrichment Samples. (B) Stacked bar plot showing the distribution of antimicrobial resistance genes (ARGs) across enrichment and environmental processing types.

### Taxonomic, Functional, and Antimicrobial Resistance Profiling of MAGs

The MAGs were classified into Actinomycetota, Bacillota, Bacteroidota, Patescibacteria, Pseudomonadota, and some unclassified phyla groups. The MAGs with a high percentage of completeness mainly belonged to phyla Bacillota and Actinomycetota. However, some MAGs from less abundant or more diverse phyla, such as Patescibacteria, show lower completeness (Figure 6C).

*Sphingomonas hankookensis* was the most dominant across all environmental sample types, particularly in Hospital samples, where it constituted more than half of the total microbial community. Other notable species included *Cutibacterium acnes*, *Methylobacterium radiotolerans*, *Pseudomonas aeruginosa*, and *E. coli*. The Aircraft Filter samples demonstrated a more diverse community structure than the Hospital samples, with a higher relative abundance of *Methylobacterium radiotolerans* and *P. aeruginosa*. *Geobacillus proteiniphilus* also showed a significant presence, particularly in Travel samples (Figure 6D).

The enrichment processing yielded a different community profile with additional species such as *Microsaccharimonas sp001897305* and *Paenibacillus humicus*. Notably, *S. hankookensis* remained dominant across all sample types, but its relative abundance was more evenly distributed with other species. *M. radiotolerans* and *C. acnes* were relatively abundant in Aircraft Filters and Hospital samples (Figure 6E).

Among the genes identified, Acyl-CoA dehydrogenase (COG1960), Acetyl-coenzyme A synthetase (COG0365), and (3S)-malyl-CoA thioesterase (COG2301) were highly prevalent across Aircraft Filter, Hospital, and Travel samples in both Enrichment and Environmental samples. In addition to these genes, Linearmycin resistance ATP-binding protein (LnrL, COG1131), a gene directly involved in antibiotic resistance, was identified in both environmental and enrichment samples. Another gene linked to virulence is the Lipopolysaccharide (LPS) export system ATP-binding protein LptB (COG1137), identified in both environmental and enrichment samples (Figure 7A).

**Figure 7:**
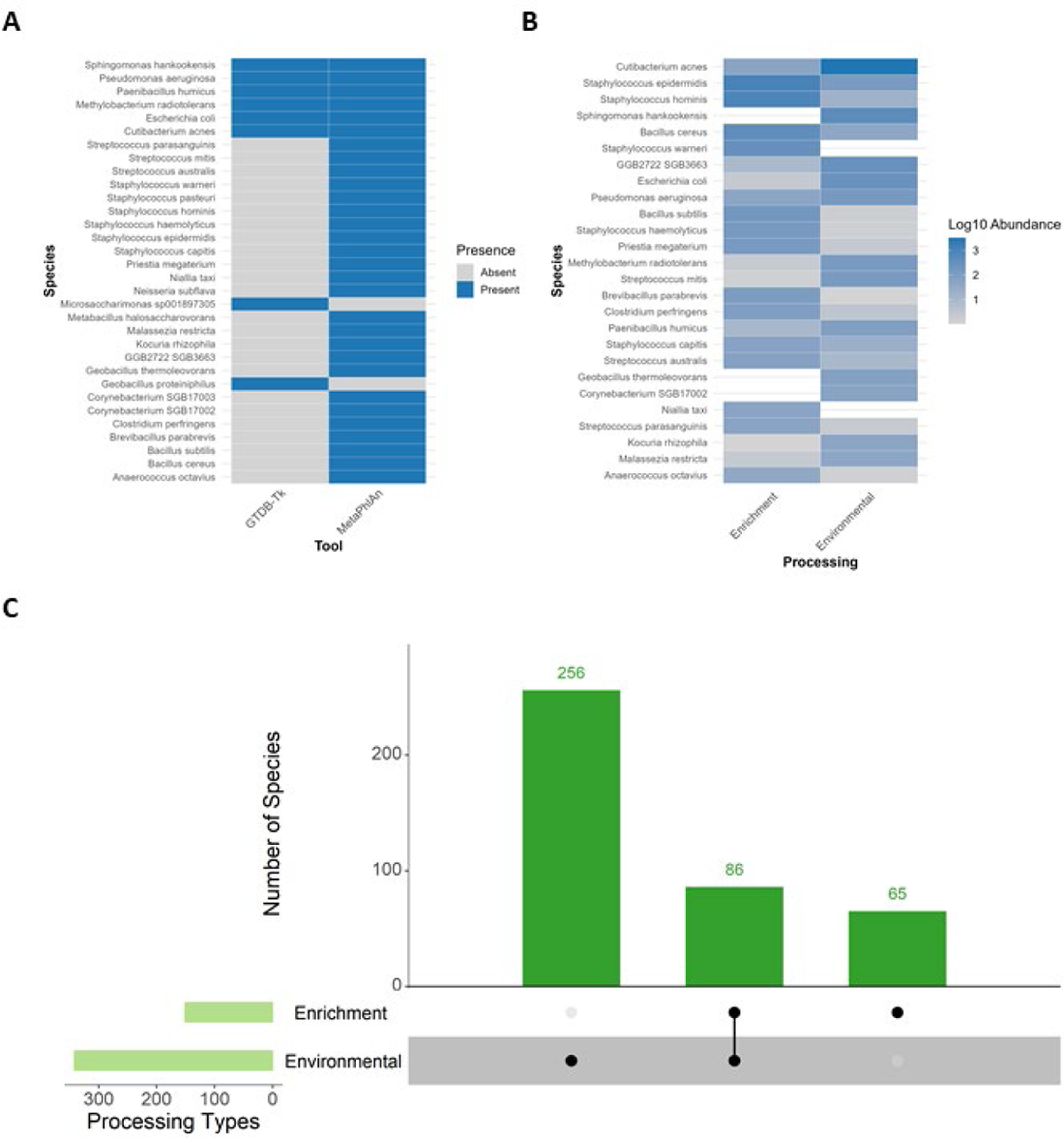
(A) The presence and absence of species were identified using two bioinformatics tools, GTDB-Tk and MetaPhlAn, and represented in a binary heatmap. (B) Log10 abundance of species detected in Enrichment and Environmental processing. (C) Comparison of microbial species diversity and relative abundance between Enrichment and Environmental processing types.

A total of 23 antimicrobial resistance types were identified in both enrichment and environmental samples. The results indicate that enrichment samples exhibited higher ARGs occurrences across various resistance categories than environmental samples. Among the most prevalent resistance types in the enrichment group were fluoroquinolone, tetracycline, penam, and cephalosporin. In contrast, the Environmental samples exhibited lower occurrences for these categories, with fluoroquinolone, tetracycline, penam, and cephalosporin being the most abundant. Furthermore, non-antibiotic antimicrobial resistances such as acridine dye and triclosan were more prevalent in the Enrichment samples (Figure 7B).

### Comparative Analysis of Microbial Diversity Across Environmental and Enrichment Processing

Across both microbial taxonomy annotation tools, MetaPhlAn and GTDB-Tk, 409 species were identified. MetaPhlAn accounted for 407 species, while GTDB-Tk identified only nine species. A total of six species, *Sphingomonas hankookens*is*, Cutibacterium acnes, Methylobacterium radiotolerans, Escherichia coli, Pseudomonas aeruginosa,* and *Paenibacillus humicus* were common to both tools. Species identified solely in GTDB-Tk (MAGs) and absent in MetaPhlAn included *Geobacillus proteiniphilus* and *Microsaccharimonas sp001897305* (Figure 8A).

Among the overall species detected, *C. acnes* was the most abundant species, consistently detected in both Enrichment and Environmental samples, along with other vital species such as *Staphylococcus epidermidis*, *Staphylococcus hominis*, *Bacillus cereus*, *E. coli*, and *P. aeruginosa*. However, certain species, including *Geobacillus thermoleovorans*, *Salmonella enterica*, and *Limosilactobacillus fermentum*, were only present in Environmental samples, while others, such as *Staphylococcus warneri* and *Niallia taxi*, were found solely in Enrichment samples (Figure 8B).

Of all the species analyzed, 256 were uniquely present in the Environmental samples, and a total of 86 species were shared between the Environmental and Enrichment categories. However, 65 species were identified solely in the Enrichment samples (Figure 11A). A statistically significant difference in species abundance was found when comparing the relative abundances of microbial species across Environmental and Enrichment processing types (Wilcoxon p-value: 1.19e-16). Environmental processing yielded higher relative abundance across species than Enrichment processing. The median abundance for species in the Environmental samples was consistently higher than in the Enrichment samples (Figure 11B).

## Discussion

Our study leveraged non-invasive air sampling using face masks and aircraft cabin filters to investigate airborne microbial communities through metagenomics. Through this approach, we aimed to provide insights into the potential of air sampling for pathogen surveillance in public and travel environments while also addressing the challenges and limitations that emerged.

Airborne microbial concentrations, estimated to range from 10⁴ to 10⁶ cells per cubic meter, are significantly lower than those found in marine or soil environments [39]. This low-biomass nature poses inherent challenges in extracting and quantifying microbial DNA [11,40]. In our study, the DNA concentrations from environmental samples remained notably low despite optimizing DNA extraction protocols. These DNA concentrations fall below the detection limits [15]. To ensure microbial DNA was present and of sufficient quality for downstream applications, we complemented our assessment with 16S rRNA gene amplification.

Enrichment cultures improved DNA yield, resulting in greater sequencing depth and enhanced the recovery of MAGs. A statistically significant difference in species abundance was observed (Wilcoxon p-value: 1.19e-16), with environmental samples showing higher relative abundance. The lower relative abundance in enrichment samples highlights the biases inherent in selective cultivation [41], which could limit the detection of broader microbial diversity. The unique species in enrichment samples (n = 65) reflects the recovery of low-abundance microbes [42,43]. Furthermore, the paired t-test (p-value: 0.5433) did not indicate a significant difference in mean species abundance between the environmental and enrichment samples. This suggests that enrichment methods may capture sufficient species to reach comparable mean abundance levels. Consequently, while enrichment techniques provide valuable insights into dominant species or targeted groups [42,44], environmental sampling appears essential to capture the natural diversity and variability of the microbial community [45,46]. Thus, combining environmental sampling with optimized enrichment protocols could reduce these biases and better capture the native diversity of airborne microbial communities.

MetaPhlAn shows high sensitivity [25] in detecting a broad spectrum of microbial species, identifying 407 species compared to GTDB-Tk’s nine. Metagenomic assembly techniques often capture only a limited portion of organisms within complex communities. This limitation is due to inadequate coverage for numerous taxa or challenges posed by genetically similar taxa that can hinder or produce erroneous assemblies [25,47] However, certain species, such as *Geobacillus proteiniphilus* and *Microsaccharimonas sp001897305*, were detected solely through MAGs, hinting at the potential for MAGs to capture species underrepresented or missed from MetaPhlAn. These findings emphasize the utility of using both short-read and MAG-based approaches to achieve a comprehensive understanding of microbial diversity.

Bacterial sequences dominated the microbial community across all sample types, while fungi comprised only a small fraction of the detected organisms. Despite including spike-in control standards containing fungal species (*Saccharomyces cerevisiae* and *Cryptococcus neoformans*), these fungi were notably absent from the sequencing data. This absence may be attributed to insufficient recovery of fungal DNA from the samples or inadequate detection of fungal sequences. Furthermore, the ZymoBIOMICS Microbial Community Standard manufacturer notes that the chemistry of the Qiagen DNeasy PowerSoil kit we utilized is not entirely compatible with the storage solution of the microbial community standard (DNA/RNA Shield). However, airborne fungi are a significant health concern worldwide, linked to various adverse effects on human health, such as asthma, hypersensitivity, and pneumonitis [48,49]. These results suggest the need for improved protocols targeting more fungal organisms.

*Cutibacterium acnes*, *Staphylococcus epidermidis*, *and Staphylococcus hominis* were consistently observed across many samples in our study. These species are commonly associated with the human skin microbiome [50,51], which suggests that human activity significantly influences microbial communities in diverse environments [50]. Previous studies have highlighted the dominance of skin-associated bacteria in indoor environments, especially in airplane cabins and hospitals [52–56]. Humans harbor 10^12^ microorganisms on their skin and 10^14^ in their digestive tract, making us a significant source of bioaerosols in indoor environments. Breathing and shedding millions of skin cells due to daily activities add to indoor environments’ microbial content [2]. In addition, face masks are often handled with the hands and can come into contact with skin and various surfaces, creating a potential pathway for skin microbial contamination [57,58].

*Cutibacterium acnes* predominates in the environmental samples. However, a notable shift in microbial community structure was observed under enrichment conditions, where *Staphylococcus spps* emerged as the dominant species across multiple sample types. This might be because the genus *Staphylococcus* is more readily cultivated than *Cutibacterium spp*., which tends to be frequently underrepresented in culture-based surveys [59,60].

The rise in antibiotic-resistant bacterial infections and the environmental dissemination of ARGs present significant challenges to public health [61]. A notable abundance of ARGs such as beta-lactamase, kanamycin kinase, and gentamicin 3-N-acetyltransferase was observed in Hospital samples. This prevalence may be related to antibiotic usage in clinical settings, which imposes selective pressure, favoring the survival and proliferation of resistant strains [62,63]. Additionally, detecting elevated levels of beta-lactamase and gentamicin 3-N-acetyltransferase in Aircraft Filters suggests that environments with high human traffic can serve as reservoirs for ARGs [64]. This finding is consistent with research demonstrating the presence of ARGs in public transportation systems, including aircraft cabins [65].

Enrichment samples display a broader range of ARGs, including resistances to fluoroquinolones, tetracyclines, penams, and cephalosporins. This broader diversity may reflect the presence of naturally occurring antimicrobial compounds that shape microbial communities. Alternatively, the enrichment conditions may have indirectly favored organisms with extensive resistance profiles, even in the absence of direct antibiotic selection. However, it is important to note that the increased number of complete MAGs from the enrichment samples may introduce a bias, potentially amplifying the observed diversity of ARGs under these conditions.

Our study identified several opportunistic pathogens [18,51,66–69], including *Staphylococcus epidermidis*, *Staphylococcus hominis*, *Escherichia coli*, and *Cutibacterium acnes*, across various sample types, particularly in high-occupancy environments such as hospitals and aircraft cabins. These species, typically commensal, can potentially cause infections, especially in immunocompromised individuals or when introduced via medical devices [66,67,69]. The presence of ARGs such as beta-lactamase and gentamicin 3-N-acetyltransferase within these pathogens is concerning, as it limits effective treatment options and can increase morbidity and mortality rates, particularly in healthcare settings where antibiotic-resistant infections pose major therapeutic challenges [70,71].

This microbial diversity underscores the potential for high-contact public environments to serve as reservoirs for ARGs and opportunistic pathogens, facilitating ARG dissemination through airborne transmission and surface contact. These findings highlight the need for ongoing surveillance of ARGs in healthcare and public transportation systems to understand better their distribution and potential transmission dynamics [72,73].

## Conclusion

Our study assesses the utility of non-invasive air sampling via face masks and aircraft cabin filters to explore airborne microbial communities in high-contact environments. Through metagenomic analysis, we identified a diverse array of microbes, including common human-associated bacteria like *Cutibacterium acnes* and *Staphylococcus epidermidis*, as well as environmental species such as *Sphingomonas hankookensis* and *Methylobacterium radiotolerans*, highlighting the impact of human activity on microbial dispersal. Addressing challenges of low biomass, we employed optimized DNA extraction and enrichment techniques, which improved DNA yield and enabled detailed microbial profiling. The presence of clinically relevant ARGs underscores the potential for hospitals and aircraft cabins to serve as ARG reservoirs. This study highlights the value of combining environmental and enrichment sampling to capture the full spectrum of microbial diversity and resistance profiles in airborne communities, and future efforts should focus on refining protocols to enhance fungal DNA recovery and integrate multiple sequencing approaches to deepen understanding of microbial diversity in these high-traffic settings.

## Supporting information

supplementary metadata

## Data Availability

All data produced are available online. Links are provided in the manuscript.

The shotgun metagenomic sequencing data generated in this study have been deposited in the National Center for Biotechnology Information (NCBI) under the BioProject accession number PRJNA1228129. The raw sequencing data, including FASTQ files, are available in the Sequence Read Archive (SRA) linked to this BioProject.

## Acknowledgments

We thank Dr. Catherine Gao for her help in obtaining face masks from clinicians and all of the clinicians and travelers who donated their used masks. This study was funded by the Walder Foundation (Grant 21-00661).

## Authors Contribution

OJ, CC and EH conceived and designed the study. OJ conducted the experiments and performed data analysis. EH, JS and AM provided feedback on the experimental design and data analysis. KT and CH provided insights on the bioinformatics outputs. OJ wrote the first draft of the manuscript, and all authors reviewed, revised, and approved the final version.

